# COVID-19 lockdowns cause global air pollution declines with implications for public health risk

**DOI:** 10.1101/2020.04.10.20060673

**Authors:** Zander S. Venter, Kristin Aunan, Sourangsu Chowdhury, Jos Lelieveld

**Affiliations:** Terrestrial Ecology Section, Norwegian Institute for Nature Research - NINA, 0349 Oslo, Norway; CICERO Center for International Climate Research, PO Box 1129 Blindern, N318 Oslo, Norway; Department of Atmospheric Chemistry, Max Planck Institute for Chemistry, 55128 Mainz, Germany; Climate and Atmosphere Research Center, The Cyprus Institute, 1645 Nicosia

**Keywords:** COVID-19, Particulate matter, Pediatric asthma, Mortality, Nitrogen dioxide, Ozone

## Abstract

The lockdown response to COVID-19 has caused an unprecedented reduction in global economic activity. We test the hypothesis that this has reduced tropospheric and ground-level air pollution concentrations using satellite data and a network of >10,000 air quality stations. After accounting for the effects of meteorological variability, we find remarkable declines in ground-level nitrogen dioxide (NO_2_: −29 % with 95% confidence interval −44% to −13%), ozone (O_3_: −11%; −20% to −2%) and fine particulate matter (PM_2.5_: −9%; −28% to 10%) during the first two weeks of lockdown (n = 27 countries). These results are largely mirrored by satellite measures of the troposphere although long-distance transport of PM_2.5_ resulted in more heterogeneous changes relative to NO_2_. Pollutant anomalies were related to short-term health outcomes using empirical exposure-response functions. We estimate that there was a net total of 7400 (340 to 14600) premature deaths and 6600 (4900 to 7900) pediatric asthma cases avoided during two weeks post-lockdown. In China and India alone, the PM_2.5_-related avoided premature mortality was 1400 (1100 to 1700) and 5300 (1000 to 11700), respectively. Assuming that the lockdown-induced deviations in pollutant concentrations are maintained for the duration of 2020, we estimate 0.78 (0.09 to 1.5) million premature deaths and 1.6 (0.8 to 2) million pediatric asthma cases could be avoided globally. While the state of global lockdown is not sustainable, these findings illustrate the potential health benefits gained from reducing “business as usual” air pollutant emissions from economic activities. Explore trends here: www.covid-19-pollution.zsv.co.za

**Significance statement:** The global response to the COVID-19 pandemic has resulted in unprecedented reductions in economic activity. We find that lockdown events have reduced air pollution levels by approximately 20% across 27 countries. The reduced air pollution levels come with a substantial health co-benefit in terms of avoided premature deaths and pediatric asthma cases that accompanied the COVID-19 containment measures.

## Introduction

In many developing nations economic growth has exacerbated air pollutant emissions with severe consequences for the environment and human health. Long-term exposure to air pollution including fine particulate matter with a diameter less than 2.5µm (PM_2.5_) and ozone (O_3_) are estimated to cause ∼8.8 million excess deaths annually (1, 2), while nitrogen dioxide (NO_2_) results in 4 million new paediatric asthma cases annually (3). Despite the apparent global air pollution “pandemic”, anthropogenic emissions remain on positive trajectories for most developing and some developed nations (4–6).

The major ambient (outdoor) air pollution sources include power generation, industry, traffic, and residential energy use (4, 7). With the rapid emergence of the novel coronavirus (COVID-19), and in particular the government enforced lockdown measures aimed at containment, economic activity has come to a near-complete standstill in many countries (8). Lockdown measures have included partial or complete closure of international borders, schools, non-essential business and in some cases restricted citizen mobility (9). The associated reduction in traffic and industry has both socio-economic and environmental impacts which are yet to be quantified. In parallel to the societal consequences of the global response to COVID-19, there is an unprecedented opportunity to estimate the short-term effects of economic activity counterfactual to “business as usual” on global air pollution and its relation to human health.

Here we test the hypothesis that reduced air pollution levels during Feb/Mar 2020 were related to the COVID-19 lockdown events. To test the hypothesis, satellite data are used to provide a global perspective over Feb/Mar, but to estimate exposure levels relevant to public health, we derive ground-level measurements from >10,000 air quality stations after accounting for meteorological variations. The air pollution anomalies during COVID-19 lockdown are then used to quantify mortality and pediatric asthma incidence that have been potentially avoided (Fig. S1). Finally we perform a counterfactual projection of the public health burden assuming NO_2_, O_3_ and PM_2.5_ anomalies during lockdown are maintained for the remainder of 2020. In doing this we do not imply that lockdown economic activity is sustainable or desirable, however, we do intend to use the current situation as an intuitive means of highlighting the significance of the often-overlooked global air pollution health crisis.

## Results and discussion

### Satellite-derived global trends

Satellite-measured tropospheric NO_2_ concentrations have decreased by an average of 10.7% (area-weighted mean with interquartile range; IQR: 32%) over inhabited areas of the globe during Feb/Mar 2020 relative to 2019 (Fig. 1A; Fig. S2). The percentage changes over areas most affected by COVID-19, including Europe and China, showed NO_2_ declines of 20% (38% IQR) and 12% (33% IQR), respectively. In contrast to NO_2_, O_3_ concentrations exhibit a net positive anomaly of +2.4% (8.4% IQR) in 2020 relative to 2019 (Fig. 1C; Fig. S2). This may be related to the emission decline of NO_x_ (=NO+NO_2_), mostly as NO, leading to reduced local titration of O_3_ (reaction of NO with O_3_). The O_3_ titration effect is relevant locally and within the planetary boundary layer, whereas further downwind photochemical O_3_ formation, with a catalytic role of NO_x_, is a more important factor. Note that lockdown impacts on NO_2_, which has an atmospheric lifetime of about a day, are clearly discernible locally, whereas those on O_3_ with a lifetime of several weeks are affected by long-distance transport associated with specific weather patterns. Further, O_3_ photochemistry in temperate latitudes during the Feb/Mar period is still slow due to low solar irradiation, whereas at lower latitudes O_3_ buildup can be significant.

**Fig. 1:**
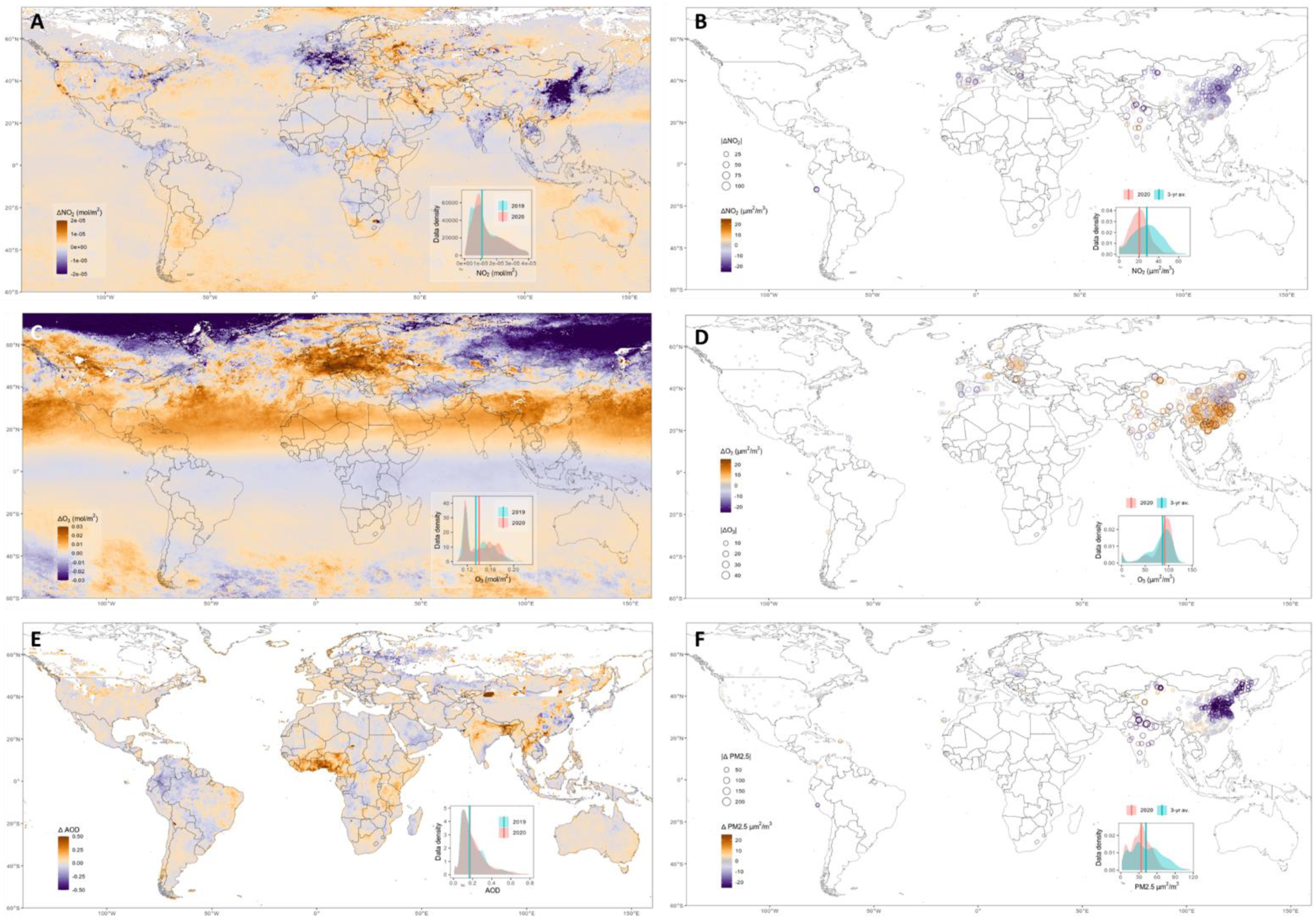
Global distribution of 2020 air pollution anomalies. Satellite and ground station measures of NO_2_ (A,B), O_3_ (C, D), aerosol optical depth (E) and PM_2.5_ (F) anomalies are mapped. Anomalies are defined as 2020 deviations from Feb/Mar 2019 average for satellite data and from Feb/Mar 3-yr averages for ground stations. Inset plots show data density distributions for anomalies over inhabited land areas.

Similarly, aerosol optical depth (AOD: a proxy for PM_2.5_) has also increased slightly (+13.2% IQR: 35%), although local declines are evident over parts of China (Fig. 1E; Fig. S2). While the lockdown impacts on NO_2_ and on ground-level O_3_ in inhabited regions are largely due to local emissions, PM_2.5_ is less locally controlled as it has an atmospheric lifetime of several days or longer in the absence of rain. For instance, European AOD levels during March 2020 were strongly influenced by dry weather with easterly winds, which carried mineral dust from West Asia, which explains some of the positive anomalies in this period. Since much of the long-distance dust transport takes place above the boundary layer (10), these AOD anomalies do not necessarily represent ground-level PM_2.5_ trends. The same is true for satellite-measured O_3_, which is strongly influenced by its generally increasing abundance above the boundary layer, especially during winter.

### Ground-level country-specific trends

While satellites provide global data coverage, they do not necessarily reflect pollutant concentrations at ground-level that are relevant to human exposure and health. Therefore we supplemented satellite data with ground-level pollutant concentrations collected by over 10,000 air quality stations. In contrast to the satellite data we used, station data allowed us to calculate a more robust 3-year baseline measure of expected pollution levels for Feb/Mar. These data largely corroborate the satellite data in that we found the same spatial patterns and net directions of Feb/Mar 2020 pollutant anomalies (Fig. 1 B, D and F). Specifically, NO_2_ declined by 22.9% (20% IQR) which equates to an absolute decline of 7.6 µg m^-3^ (9 µg m^-3^ IQR). O_3_ increased by 5.4% (18% IQR) whereas particulate matter (PM_2.5_) declined by 17.2% (30% IQR). The direction and magnitude of PM_2.5_ change near the surface is different to the AOD measured by satellites, highlighting the importance of ground-level measurements to complement satellite-derived global trends.

Focusing on the ground-level trends is illustrative of the change at both global (Fig. 2A-C) and country (Fig. 2 D - F; Fig. S3) scales. Here, the deviation in NO_2_ and PM_2.5_ levels from 3-yr average values increases significantly from mid-Jan onward (Fig. 2). The timing of the initial deviation is potentially an effect of the dramatic air pollution reductions in China (Fig. 2D; Fig. S4) coincident with the rapid lockdown response in Wuhan province at the outset of COVID-19. Thereafter, the spread of COVID-19 led to lockdowns in various countries, associated with a greater negative deviation in NO_2_ and PM_2.5_ from 3-yr baseline values (Fig. 2). Some notable outliers include Australia and Mexico. Australia exhibited drastic declines in PM_2.5_ from January onward likely reflecting the tail-end of the recent wildfires (11). The rapid decline in NO_2_ over Mexican stations is more difficult to explain, particularly given that Mexico, along with Taiwan, Slovakia and Sweden, was one of the few countries not to enforce any national lockdown measures.

**Fig. 2:**
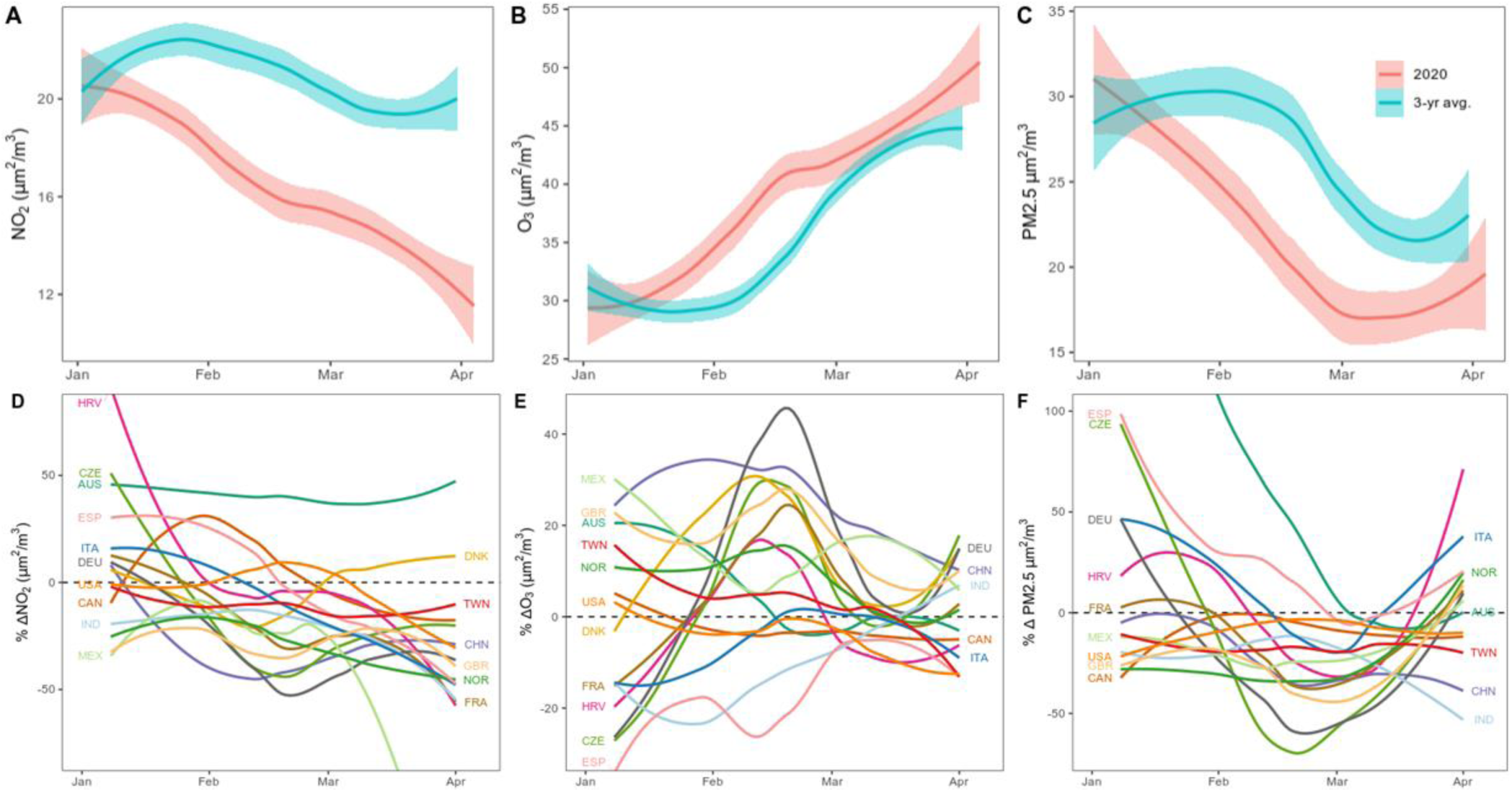
Ground-level air pollution time series. Weekly time series for ground station pollutant concentrations are plotted for Feb/Mar 2020 and the 3-yr average for the equivalent weeks (A, B, C). Loess regression lines and 95% confidence interval ribbons show globally averaged trends (n = 30 countries). Country-specific time series showing percentage deviation from long-term means are plotted in D, E and F. For country code reference refer to: www.iso.org/obp/ui/

The trends for O_3_ and PM_2.5_ are more heterogeneous over space (Fig. 1D, F) and time (Fig. 2E, F) relative to the ubiquitous declines in NO_2_. For instance, increases in O_3_ over southern China differ significantly from the decreases observed over the Wuhan province, the epicenter of COVID-19 (Fig. 1D). We expect this to be a consequence of synoptic redistribution of O_3_ by atmospheric circulations. Similarly, the local decreases over parts of Spain are in contrast to increases observed over eastern Europe. This is not surprising given that O_3_ is affected by long-distance transport as well as non-linear chemical interactions with volatile organic compounds (VOCs) and NO_x_, mediated by mesoscale and urban canopy weather patterns (12).

### Direct links to COVID-19 lockdown and health outcomes

To test our primary hypothesis that pollution anomalies were directly associated with COVID-19 lockdown events, we calculated average ground-level concentrations for each country separately. Instead of averaging over Feb/Mar, we focus on the two weeks after lockdowns were announced in each country. We first corrected for the effects of local and meso-scale weather patterns (temperature, humidity, precipitation and wind speed) which can significantly affect ground-level pollutant concentrations (13, 14) and thereby compromise any observable effect of COVID-19 lockdowns. Using regression models, we estimated the lockdown-attributable anomaly (Fig. S1) as the difference between observed and expected pollutant concentrations given weather during lockdown.

We found a net decline of about 20% (5% to 35% - 95% confidence interval) across all three pollutants in countries where significant anomalies were detected. There were significant declines in NO_2_ (29 %; 13% to 44%) and O_3_ (11%; 2% to 20%), however our model estimates were not able to control for the confounding effects of weather enough to detect significant declines in PM_2.5_ (9% decline; −10 to 28%; Fig. S5). Indeed, our meteorological-control models were able to explain less of the variance in PM_2.5_ (R^2^ = 0.45) compared to NO_2_ (R^2^ = 0.54) and O_3_ (R^2^ = 0.72; Table S1). This suggests PM_2.5_ has a weaker coupling to land-transportation and small business activity declines during lockdown compared to NO_2_ and O_3_. In Many countries PM_2.5_ is more strongly linked to residential energy use, power generation and agriculture (7). In addition, PM_2.5_ is significantly influenced by long-distance atmospheric transport of mineral dust and therefore the local effects of economic activity may be diluted or even overwhelmed (15).

Using the two week post-lockdown anomalies in combination with published exposure-response functions for NO_2_ (16, 17), O_3_ (17, 18) and PM_2.5_ (17, 19), we estimated changes in daily all-cause mortality burden and pediatric asthma incidence. During the two weeks post-lockdown, there were a total of 7400 (340 to 14600) deaths and 6600 (4900 to 7900) pediatric asthma cases avoided across 27 countries with recorded COVID-19 mitigation measures (Fig. 3; Table S2). The number of PM_2.5_-related deaths avoided (6800; 60 to 13700) exceeded those related to NO_2_ (540; 300 to 800) and O_3_ (50; 10 to 80). While for pediatric asthma incidence, NO_2_ reductions contributed to more avoided cases (5700; 4500 to 6800) compared to O_3_ (50; 40 to 60) and PM_2.5_ (850; 300 to 1000). In China and India alone, the PM_2.5_-related reductions in mortality burden were 1400 (1100 to 1700) and 5300 (1000 to 11700), respectively (Fig. 3C). These are countries with both the highest baseline pollution levels and population densities, and therefore have the most to gain from pollutant declines.

**Fig. 3:**
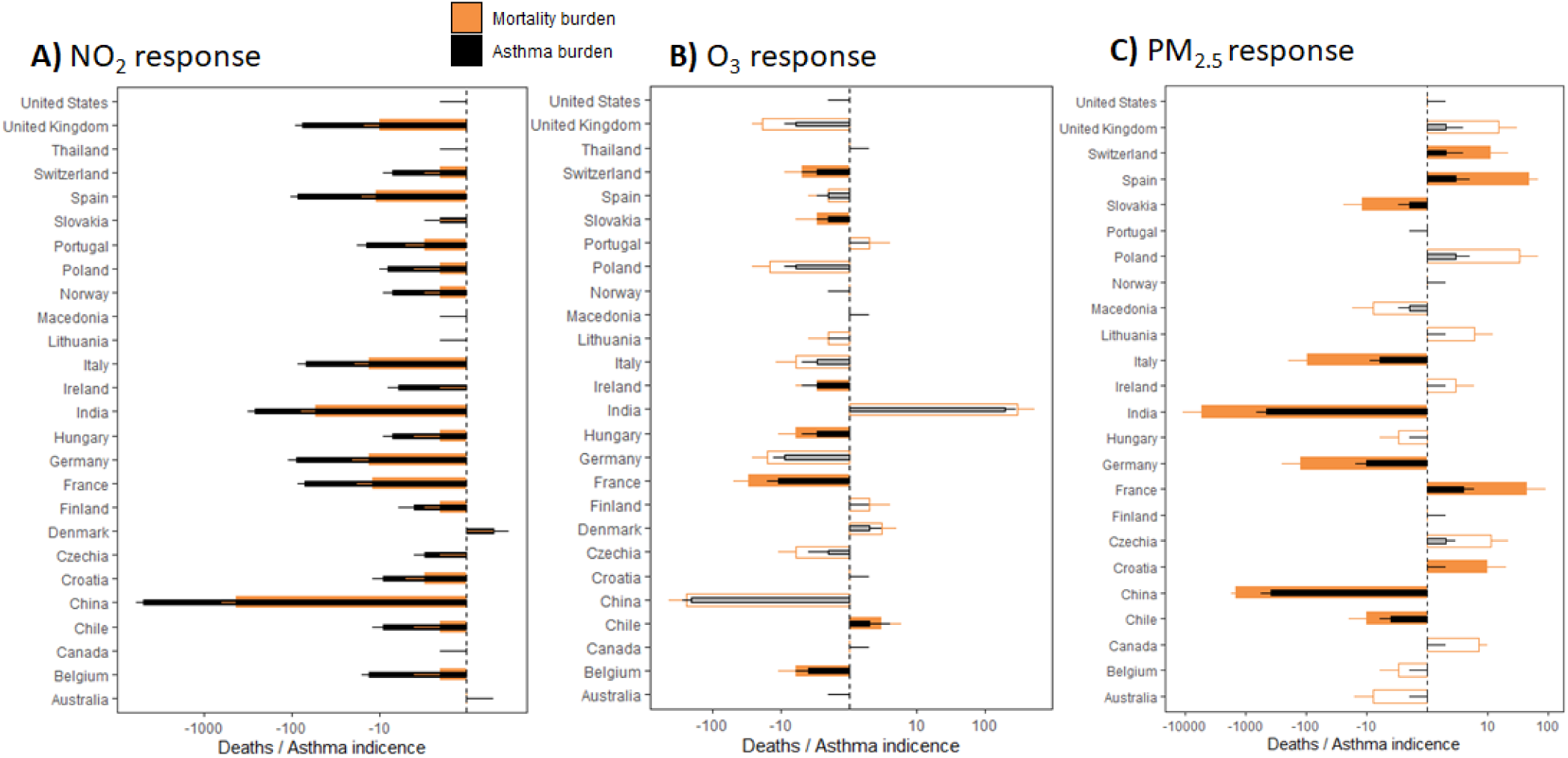
Post-lockdown health burden changes attributable to air pollution. Air pollution anomalies during two weeks post-lockdown are converted to mortality and asthma responses (n = 27 countries). Total health burden avoided (-ve) and incurred (+ve) values are presented with bars along a log-transformed x-axis. 95% uncertainty intervals are marked with error bars. Hollow bars represent estimates where the change in pollutant concentrations were not significant (p > 0.05) after accounting for weather variations (Fig. S5).

Furthermore, we performed a counterfactual projection of reduced health burden assuming ground-level air pollution deviations experienced during lockdown (Fig. S5) are maintained for the remainder of 2020 (Apr-Dec). The cumulative effect of the reduction in NO_2_, O_3_ and PM_2.5_ over the remainder of 2020 is that 0.78 (0.09 to 1.5) million deaths and 1.6 (0.8 to 2) million pediatric asthma cases could be avoided (Fig. 4; Table S3). Our findings suggest that, in spite of the modest response of PM_2.5_, countries would have much to gain in maintaining PM_2.5_ lockdown levels because that would prevent 0.6 (0.01 to 1.3) million deaths and 1.1 (0.4 to 1.4) million pediatric asthma cases which is 3- and 5-fold higher than those from NO_2_ and 5- and 30-fold higher than those from O_3_ (Fig. 4). The bulk of the benefit gained would take place during the latter half of the year when air pollution levels are at their highest over countries with the largest air pollution health burden (i.e. India and China).

**Fig. 4:**
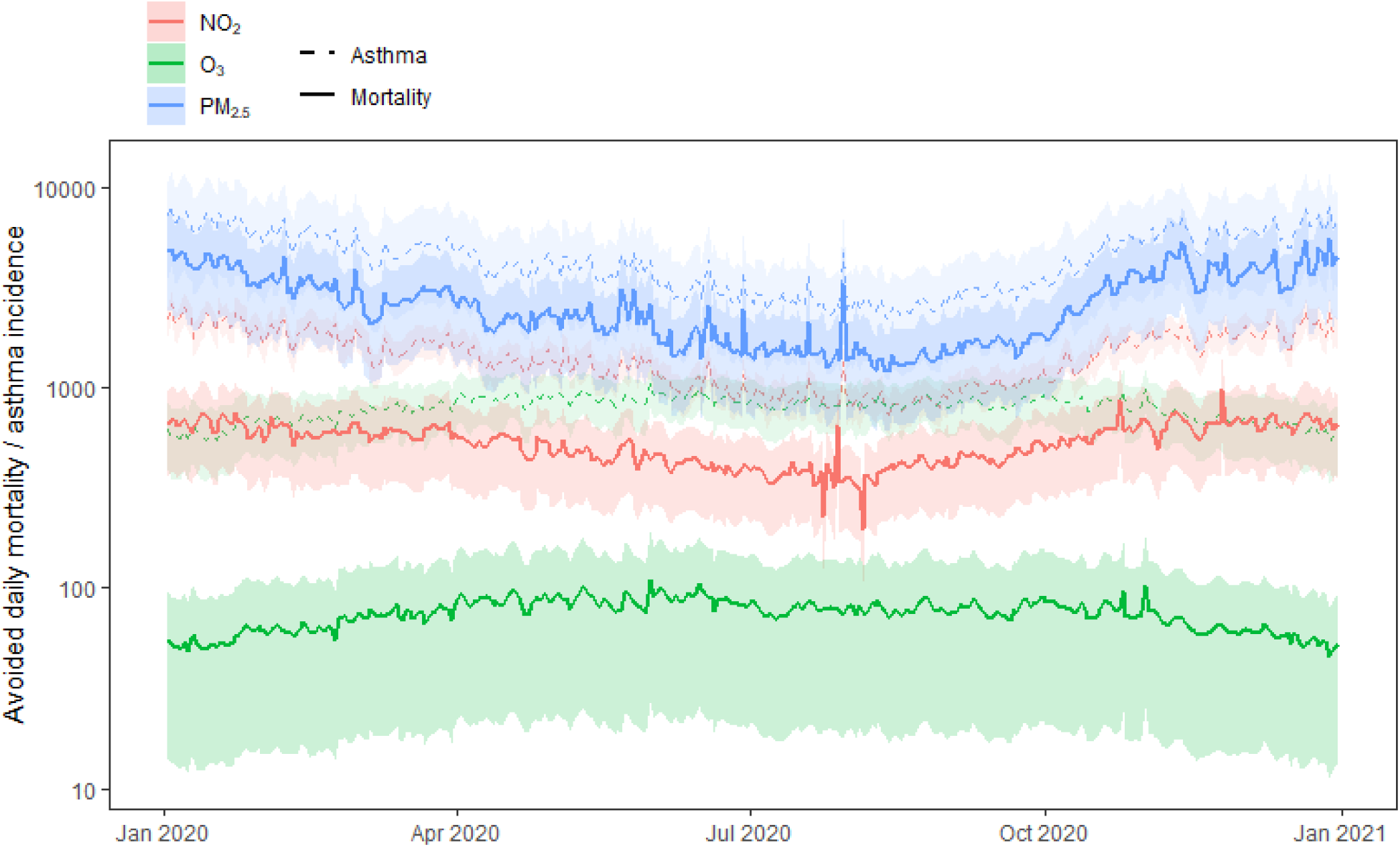
Projected daily health outcomes over 2020. Potential daily premature deaths (solid lines) and asthma incidence (dashed lines) that might be avoided assuming pollutant levels remain at lockdown levels (NO_2_: −29%; O_3_: −11%; PM_2.5_: −9%). Lines reflect global averages (n = 27 countries) with 95% confidence interval ribbons.

### Limitations and perspectives

Making explicit links between ambient air pollution and human health burden relies on several assumptions that are difficult to verify *apriori*. First, using relative risk rates from select meta-analysis (17) and multi-city (n>406) short-term time-series association studies (18, 19) to make inference over entire countries rests on the assumption that city- or cohort-specific response rates are generalizable to broader populations. While this is likely to introduce uncertainty, the dearth of representative data necessities these generalizations, and this approach has been used by numerous studies at the global scale (2, 3). Further, we acknowledge that our results are affected by harvesting effects, where premature deaths attributed to air pollution might have occurred in the immediate future (20). Note that this also applies to death counts attributed to COVID-19. We also acknowledge that we do not account for indoor sources of PM_2.5_ pollution which are unlikely to be reduced by lockdown measures. As smoke from household stoves add substantially to population exposure for people dependent on solid fuels, accounting for ambient air pollution only could imply a misclassification of exposure and biased health burden estimates (21). Finally, the baseline mortality rates we use are from 2017 (22) and therefore may be prone to ignoring before and after COVID-19 onset differences in baseline mortality incidence.

Despite these assumptions and the associated uncertainty, the analysis and results presented here can provide useful insights to raise awareness and orientate interventions regarding the global effects of air pollution on human health. They should be interpreted as preliminary lessons from the Corona crisis. As the science evolves, and the COVID-19 pandemic plays out, empirical data will emerge to fill in the knowledge gaps and uncertainties associated with air pollution health burden attribution. It is expected that the two-week lockdown effects calculated here will be an underestimate of the full effect because most lockdowns will likely last much longer than two weeks. Further, we were not able to calculate the extent to which air pollution reductions have mitigated COVID-19 deaths. For instance, positive associations have been reported between air pollution and SARS case fatalities in China during 2003 (23) and preliminary analysis has revealed similar patterns for COVID-19 (24, 25). Therefore our estimates may represent lower limits after considering the air pollution reductions as a cofactor in COVID-19 case recoveries.

## Conclusions

Reducing economic activity to levels equivalent to a lockdown state are impractical, yet maintaining “business as usual” clearly exacerbates global pollutant emissions and associated deaths. Our study documents the dramatic short-term effect of global reductions in transport and economic activity on reducing ground-level NO_2_, with mixed effects on O_3_ and PM_2.5_ concentrations. Maintaining reductions in pollutant emissions corresponding to lockdown conditions can substantially reduce the global burden of disease. We by no means imply that global pandemics such as the COVID-19, nor lockdown actions, are beneficial for public health. However, we suggest the current situation is a useful lens through which to view the global air pollution “pandemic”. Time will tell how significant the change in health burden has actually been. Nevertheless, the early evidence presented here suggests it is likely significant. Reduced premature mortality from air pollution thus appears as a co-benefit of the minimized number of deaths from the lockdown measures, although more accurate, quantitative assessments must await termination of the crisis. Finding economically and socially sustainable alternatives to fossil fuel based transport and industry are another means of reaching the pollutant declines we have observed during the global response to COVID-19.

## Materials and methods

In brief, the methodological workflow (Fig. S1) described below involves collecting satellite and in-situ air pollution time series data to estimate anomalies during the 2020 COVD-19 period relative to different baseline levels. Regression models are used to correct for the potential effects of weather-related variations on pollutant levels during lockdown. The resulting estimates of pollutant anomalies are related to established health burden estimates for short-term premature mortality and pediatric asthma incidence attributable to air pollution. The sample of countries used in each step varies dependent on the data availability. Results for satellite data contain all countries (n = 196). For ground-station anomalies there were 30 countries in total, however lockdown anomalies and health burden statistics are only reported for those with recorded lockdown measures (n = 27).

### Satellite data

All remote sensing data analyses were conducted in the Google Earth Engine platform for geospatial analysis and cloud computing (26). All data was extracted at a global scale and aggregated to the mean for each country. Data outside of inhabited areas (ocean, freshwater, desert etc.) were excluded from the analysis using the Global Human Settlement Layer produced by the European Joint Research Centre which defines inhabited rural and urban terrestrial areas (27). We did this because our main hypothesis was linked to human exposure and therefore we aimed at pollution measures that were relevant to inhabited land surfaces.

We collected nitrogen dioxide (NO_2_) and ozone (O_3_) data from the TROPOspheric Monitoring Instrument (TROPOMI), on-board the Sentinel-5 Precursor satellite (28). TROPOMI has delivered calibrated data since July 2018 from its nadir-viewing spectrometer measuring reflected sunlight in the visible, near-infrared, ultraviolet, and shortwave infrared. Recent work has shown that TROPOMI measurements are well correlated to ground measures of NO_2_ (29, 30) and O_3_ (31). We filtered out pixels that are fully or partially covered by clouds using 0.3 as a cutoff for the radiative cloud fraction. As a proxy for atmospheric fine particulate matter (PM_2.5_), we collected aerosol optical depth (AOD) data from the cloud-masked MCD19A2.006 Terra and Aqua MAIAC collection (32). This dataset has been successfully used to map ground-level PM_2.5_ concentrations (33, 34). Global median composite images for NO_2_, O_3_ and AOD were then calculated for the months of February and March 2019 and 2020.

### In-situ data

Although satellite data have the advantage of wall-to-wall global coverage, there are some drawbacks: (1) TROPOMI does not extend back far enough to obtain an adequate baseline measure with which to compare 2020 concentrations; (2) MODIS and TROPOMI collect information within either the total (O_3_ and AOD) or tropospheric (NO_2_) column which do not necessarily reflect pollutant levels experienced on the ground. Therefore, we also collected NO_2_, O_3_ and PM_2.5_ data from >10,000 in-situ air quality monitoring stations to supplement the satellite data. These data were accessed from the OpenAQ Platform and originate from government- and research-grade sources. See www.openaq.org for a list of sources. Despite the reliability of the sources, we inspected pollutant time series for each country and removed spurious outliers in the data with z-scores exceeding an absolute value of 3. Following quality control, we were left with data representing 30 countries.

### Quantifying air pollution anomalies

We used two approaches to quantify air pollution anomalies coincident with COVID-19 during Feb/Mar 2020. We refer to these as (1) the Feb/Mar differential, and (2) the lockdown differential (Fig. S1). For the Feb/Mar differential we calculated average pollutant levels for Feb/Mar each year between 2017 and 2020. The differential was defined as the difference between 2020 values and the average of those for a 3-year baseline (2017-2019). For satellite data the baseline was the 2019 Feb/Mar average due to limited temporal extent of TROPOMI data, however for ground-stations we considered a 3-year (2017-2019) average for the Feb/Mar period.

Air pollution anomalies measured with the Feb/Mar differential approach may smooth over the effect of COVID-19 given that country-specific lockdowns or mitigation actions occurred at different times. For instance China went into lockdown in Jan/Feb whereas the majority of lockdowns in other countries occured in March. Therefore we attempted to isolate the effect of COVID-19 mitigation measures by calculating lockdown pollutant levels for each country separately. We searched online media and news articles to identify the starting date of lockdown for each country. Sources were cross-referenced to account for erroneous reporting. We defined two levels of lockdown intensity including moderate and severe lockdowns. Moderate lockdowns involved partial or full closure of borders and flights, government advisories for citizens to work from home, closure of schools, and limiting gathering sizes. Severe lockdowns included government-enforced movement restrictions or curfews and closure of all non-essential businesses. This resulted in a sample of 27 countries that reported lockdown measures and which we had ground-level air pollution data for.

Air pollution anomalies measured during two weeks post-lockdown are not necessarily attributable to reduced economic activity, but may be an artifact of meteorological variability coincident with the onset of COVID-19. Therefore we adopted a modelled differential approach to correct for the effect of meteorological parameters on air pollution trends. This involved developing a model based on historical data to estimate what the expected air pollution levels for 2020 lockdown dates should have been given the prevailing weather conditions and time of year. We performed multiple linear regression of weekly pollutant concentrations on temperature, humidity, precipitation and wind speed derived from the Global Forecast System (GFS) of the National Centers for Environmental Prediction (NCEP) between Jan 2017 and Apr 2020. We accounted for the effect of seasonal fluctuations and long-term trends by including month and year as fixed effects in the model. We calculated the sin and cos component of the month variable to account for its cyclical nature. Using models trained on historical data, we predicted the expected pollutant levels for the two lockdown weeks. The modelled differential is then the difference between this predicted value and the observed pollutant concentrations during lockdown (Fig. S1). This differential has been attributed to COVID-19 mitigation measures with greater confidence than simple comparisons with 3-yr baseline values.

### Linking air pollution anomalies to public health burden

To relate COVID-19 lockdown air pollution anomalies to all-cause mortality and pediatric asthma incidence we applied short-term (daily) exposure-response relationships reported in recent literature. We obtained relative risks from recent studies on the relationship between daily mortality and O_3_ (18) and PM_2.5_ (19) resulting from the Multi-City Multi-Country (MCC) Collaborative Research Network (35). For NO_2_-mortality responses, we used relative risks reported in a meta-analysis which controlled for the effect of particulate matter to extract excess mortality solely attributable to NO_2_ (16). Pediatric (< 18 years) short-term relative risks for asthma incidence in response to NO_2_, O_3_ and PM_2.5_ were derived from a global meta-analysis of 87 studies (17). These data are not country-specific and we therefore applied the same relative risk rate to all countries in our study.

Daily health burden (premature mortality and asthma incidence) for each country was derived with the formula:

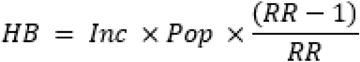

Where *Inc* is the baseline mortality or asthma incidence rate and *Pop* is the total population. *Inc* for mortality and asthma were obtained from the Institute for Health Metrics and Evaluation (IHME) for the 27 countries in our study (22), downloadable at the GDBx platform (http://ghdx.healthdata.org/). Population estimates for 2020 were calculated using the Gridded Population of the World (GPWv14) dataset (36). *RR* is the relative risk derived from the literature after log-linear transformation. We used log-linear transformation as adopted by many others (3, 37) to prevent assumptions of linearity in the relationship between pollutant concentrations and health outcome. We derive the transformed *RR* using:

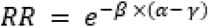

where αis the pollutant concentration and *γ* is the low concentration threshold below which there is no risk of mortality or asthma incidence. Low concentration thresholds were derived from the associated literature for O_3_ at 70 µg m^-3^ (18); PM_2.5_ at 4.1 µg m^-3^ (19) and NO_2_ at 2 ppb (3). Here ***β*** is defined by the function:

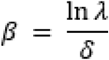

where ƛ is the relative risk reported in the literature and δ is the concentration increment used. All three studies reported results relative to a of 10 µg m^-3^.

The air pollution health burden anomaly coincident with COVID-19 lockdown was defined as:

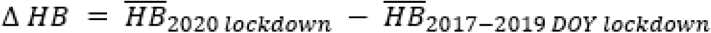

Where *DOY* is the days-of-year equivalent for each country’s two weeks lockdown dates. We use 95% confidence intervals reported in the literature to derive error margins around our change estimates. Health burden estimates are made for each day during lockdown events during 2020 and the past three years for comparison. We also perform a counterfactual forecasting assessment for 2020 where we assume the lockdown reductions in NO_2_, O_3_, PM_2.5_ are sustained for the remainder of the year. Using the resulting daily forecasts we calculated the total avoidable air pollution related mortalities and new asthma incidence.

## Data Availability

Data will be made available upon request.

## Supplementary tables and figures

**Fig. S1:**
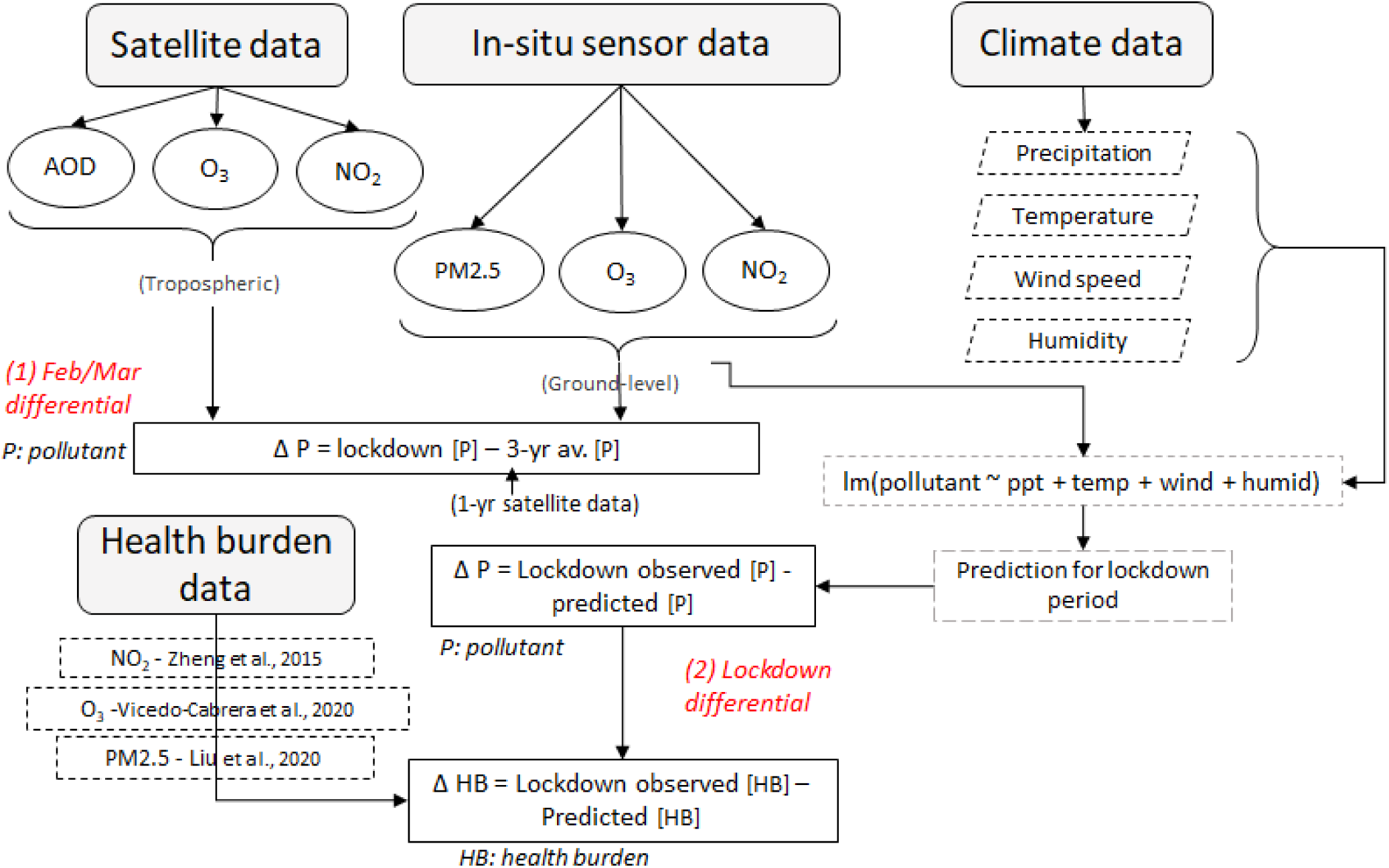
Methodological workflow for paper. Two types of air pollution (*P*) anomaly are calculated including *Feb/Mar differential* and *Lockdown differential*. The first is the difference between the Feb/Mar 2020 and the average for the same days during the previous three years (2017-2019; ground-station data) or one year (satellite data). The *Lockdown differential* is the difference between observed and predicted pollutant levels for two weeks post-lockdown. Predictions are made to account for the confounding effects of weather variability using a regression model. These differentials are used to calculate the change in mortality or asthma burden (*HB*) as a result of COVID-19 induced pollution anomalies. Relative risk rate functions are extracted from the literature outlined with dashed lines (refer to reference list in main manuscript for full references).

**Fig. S2:**
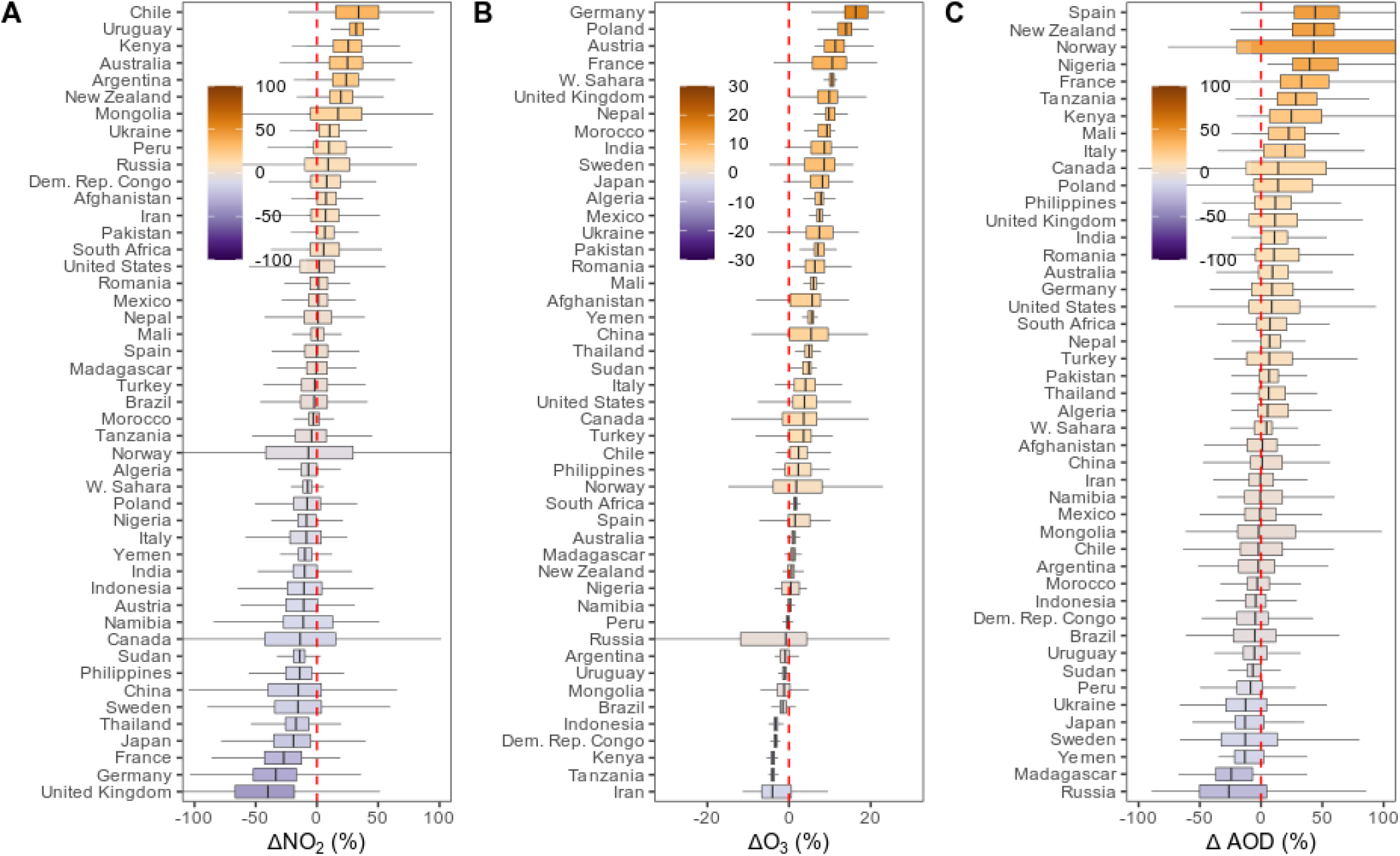
Satellite-derived air pollution Feb/Mar anomalies. Percentage temporal differentials (Feb/Mar 2020 vs Feb/Mar 2019) in atmospheric NO_2_, O_3_ and aerosol optical depth (AOD) per country. Box and whisker plots show the spread of the data (each data point is a satellite pixel within a country) around the median value.

**Fig. S3.**
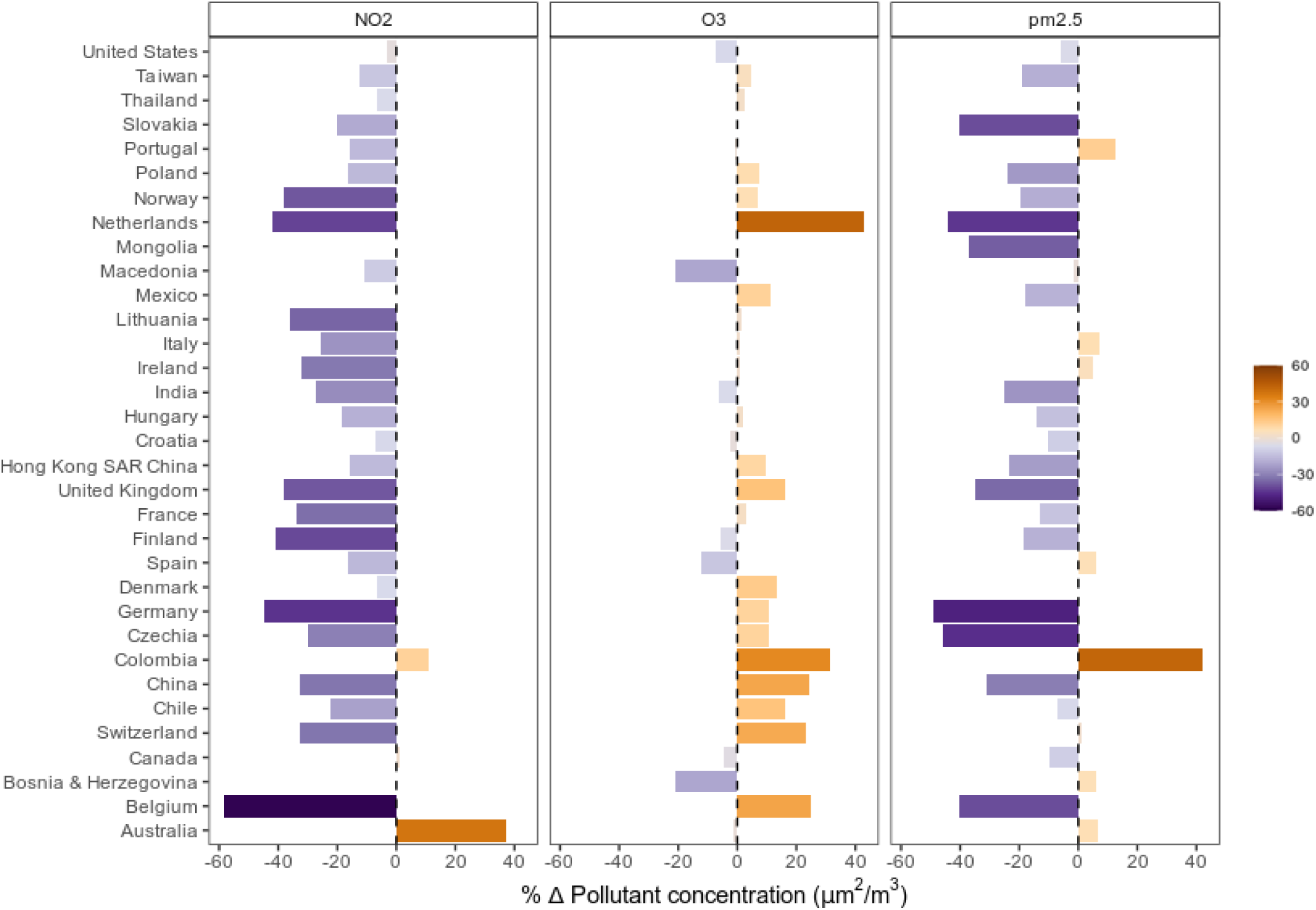
Ground-level air pollution Feb/Mar anomalies. Percentage Feb/Mar differentials (Feb/Mar 2020 vs 3-yr average for Feb/Mar) in atmospheric NO_2_, O_3_ and PM_2.5_ per country with air quality station data. Anomalies are expressed as percentage differences with bars.

**Fig. S4.**
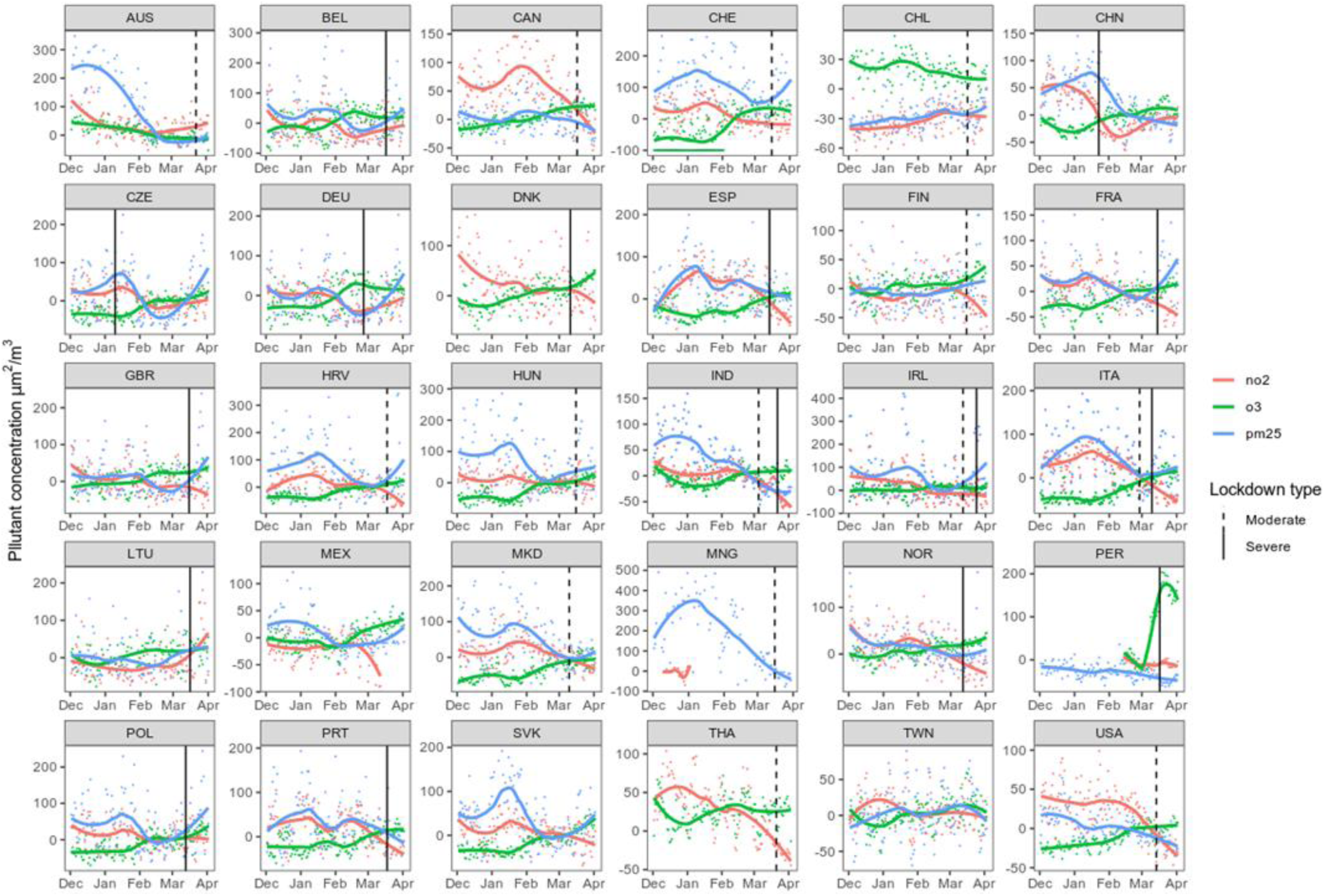
Pollutant time series and lockdown dates. Daily time series of ground-level NO_2_, O_3_ and PM_2.5_ per country with dates of lockdown indicated by vertical lines. Smoothed loess regression lines are fitted to indicate moving averages. For country code reference refer to: www.iso.org/obp/ui/

**Fig. S5.**
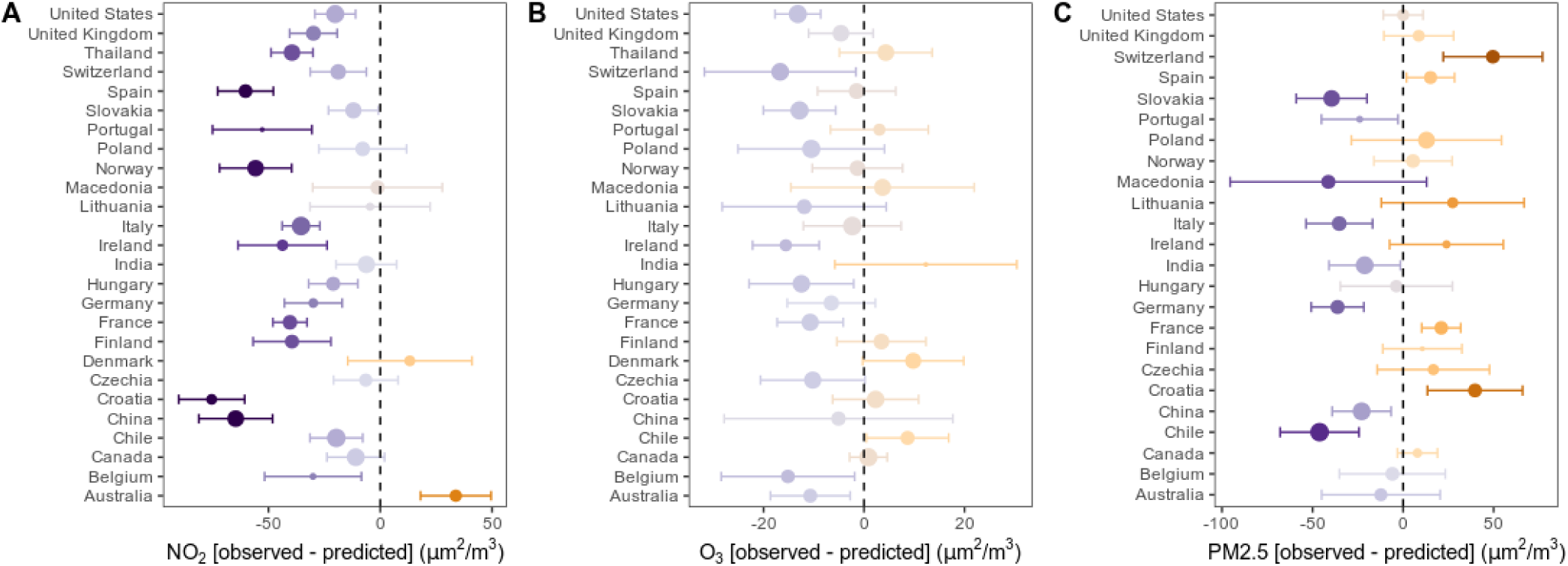
Ground-level air pollution lockdown anomalies corrected for weather variations. Percentage lockdown differentials (observed vs predicted concentrations for lockdown dates) in atmospheric NO_2_, O_3_ and PM_2.5_ per country with air quality station data. Anomalies are expressed as percentage differences with points and 95% confidence intervals with error bars. Predicted values are based on regression models that account for the effects of weather variations during lockdown. Points are sized relative to the R^2^ of the model ranging from 0.2 to 0.9.

**Table S1.** Regression model performance. Air pollutant concentrations were regressed on meteorological variables (temperature, humidity, precipitation and wind speed) to predict what air pollutant concentrations were expected to be during lockdown dates. Separate models were built for each country and the resulting *R*^*2*^ and *p-values* are presented.

|  | **NO<sub>2</sub>** |  | **O<sub>3</sub>** |  | **PM<sub>2.5</sub>** |  |
| --- | --- | --- | --- | --- | --- | --- |
|  | ***R*<sup>2</sup>** | ***p*-value** | ***R*<sup>2</sup>** | ***p*-value** | ***R*<sup>2</sup>** | ***p*-value** |
| Australia | 0.432 | 7.4E-13 | 0.598 | 6.0E-26 | 0.412 | 1.5E-13 |
| Belgium | 0.279 | 3.1E-05 | 0.583 | 4.9E-21 | 0.452 | 2.4E-13 |
| Canada | 0.775 | 8.4E-40 | 0.869 | 1.0E-60 | 0.278 | 5.3E-06 |
| Chile | 0.835 | 7.9E-50 | 0.601 | 1.1E-24 | 0.786 | 9.5E-45 |
| China | 0.682 | 3.9E-21 | 0.620 | 4.5E-19 | 0.730 | 7.4E-34 |
| Croatia | 0.372 | 3.7E-09 | 0.815 | 8.7E-49 | 0.453 | 1.4E-14 |
| Czechia | 0.472 | 4.5E-14 | 0.799 | 1.9E-46 | 0.332 | 8.4E-09 |
| Denmark | 0.376 | 2.1E-08 | 0.714 | 1.7E-29 | 0.446 | 5.2E-05 |
| Finland | 0.515 | 1.1E-16 | 0.696 | 2.0E-33 | 0.209 | 2.4E-04 |
| France | 0.547 | 3.2E-18 | 0.799 | 9.8E-46 | 0.437 | 1.1E-13 |
| Germany | 0.338 | 1.6E-07 | 0.672 | 7.6E-30 | 0.468 | 3.6E-15 |
| Hungary | 0.527 | 6.0E-17 | 0.822 | 1.1E-49 | 0.385 | 5.1E-11 |
| India | 0.737 | 5.5E-25 | 0.367 | 2.8E-07 | 0.725 | 2.4E-35 |

|  | NO<sub>2</sub> |  | O<sub>3</sub> |  | PM<sub>2.5</sub> |  |
| --- | --- | --- | --- | --- | --- | --- |
|  | *R*<sup>2</sup> | *p-value* | *R*<sup>2</sup> | *p-value* | *R*<sup>2</sup> | *p-value* |
| Ireland | 0.394 | 2.1E-09 | 0.519 | 2.1E-17 | 0.245 | 4.1E-04 |
| Italy | 0.829 | 3.5E-40 | 0.897 | 1.8E-57 | 0.505 | 2.8E-14 |
| Lithuania | 0.301 | 3.7E-05 | 0.640 | 2.0E-23 | 0.325 | 4.1E-07 |
| Macedonia | 0.522 | 5.6E-17 | 0.781 | 1.2E-43 | 0.464 | 3.7E-10 |
| Norway | 0.668 | 6.0E-29 | 0.686 | 2.6E-33 | 0.455 | 1.6E-15 |
| Peru | 0.450 | 2.2E-03 | 0.820 | 2.9E-17 | 0.406 | 6.7E-10 |
| Poland | 0.549 | 7.5E-07 | 0.870 | 1.9E-25 | 0.653 | 1.1E-11 |
| Portugal | 0.266 | 3.6E-04 | 0.513 | 2.3E-15 | 0.223 | 6.6E-04 |
| Slovakia | 0.648 | 6.7E-22 | 0.866 | 4.8E-51 | 0.626 | 1.3E-22 |
| Spain | 0.503 | 8.4E-16 | 0.693 | 7.8E-33 | 0.416 | 7.5E-13 |
| Switzerland | 0.605 | 4.4E-16 | 0.829 | 2.4E-38 | 0.446 | 2.3E-10 |
| Thailand | 0.670 | 1.0E-28 | 0.757 | 3.6E-41 | 0.453 | 5.7E-05 |
| United Kingdom | 0.569 | 8.4E-21 | 0.750 | 7.5E-41 | 0.360 | 1.8E-10 |
| United States | 0.795 | 1.2E-42 | 0.865 | 3.4E-59 | 0.368 | 1.2E-09 |

**Table S2.** Lockdown health burden response. Pollutant-related mortality and pediatric asthma cases avoided for each country during two weeks of lockdown. Country averages and 95% confidence intervals are reported with negative (-) signs representing cases where health burden has increased. Numbers are rounded to the nearest whole number. Values with significant (p < 0.05) pollutant anomalies after correcting for meteorological parameters are indicated with *.

| Country | Mortality |  |  | Asthma |  |  |
| --- | --- | --- | --- | --- | --- | --- |
|  | NO<sub>2</sub> | O<sub>3</sub> | PM<sub>2.5</sub> | NO<sub>2</sub> | O<sub>3</sub> | PM<sub>2.5</sub> |
| Australia | 0 [0; 0]* | 0 [0; 0]* | 7 [-1; 14] | 0 [0; 0]* | 0 [0; 0]* | 0 [0; 0] |
| Belgium | 1 [1; 2]* | 5 [1; 9]* | 2 [0; 5] | 12 [9; 14]* | 3 [2; 4]* | 0 [0; 0] |
| Canada | 0 [0; 0] | 0 [0; 0] | -6 [-4; -8] | 0 [0; 0] | 0 [0; 0] | 0 [0; 0] |
| Chile | 1 [1; 2]* | -2 [-1; -4]* | 9 [-23; 42]* | 8 [6; 9]* | -1 [-1; -2]* | 3 [1; 3]* |
| China | 427 [235; 619]* | 247 [35; 440] | 1444 [1127; 1761]* | 4992 [3973; 5962]* | 212 [127; 265] | 381 [143; 478]* |
| Croatia | 2 [1; 2]* | 0 [0; -1] | -9 [2; -21]* | 8 [6; 10]* | 0 [0; 0] | 0 [0; -1]* |
| Czechia | 0 [0; 0] | 5 [1; 9] | -10 [2; -23] | 2 [1; 2] | 1 [1; 2] | -1 [0; -1] |
| Denmark | 0 [0; 0] | -2 [-1; -3] | 0 [0; 0] | -1 [-1; -1] | -1 [0; -1] | 0 [0; 0] |
| Finland | 1 [0; 1]* | -1 [0; -1] | 0 [1; -1] | 3 [2; 3]* | 0 [0; 0] | 0 [0; 0] |
| France | 11 [6; 16]* | 29 [9; 47]* | -44 [8; -97]* | 69 [55; 84]* | 10 [6; 14]* | -3 [-1; -4]* |
| Germany | 12 [7; 18]* | 15 [5; 25] | 125 [-23; 276]* | 88 [70; 106]* | 8 [4; 10] | 9 [3; 11]* |
| Hungary | 1 [1; 2]* | 5 [1; 8]* | 2 [0; 5] | 6 [5; 7]* | 2 [1; 2]* | 0 [0; 0] |
|  | NO<sub>2</sub> | O<sub>3</sub> | PM<sub>2.5</sub> | NO<sub>2</sub> | O<sub>3</sub> | PM<sub>2.5</sub> |
| India | 52 [29; 76] | -300 [-77; -522] | 5313 [998; 11763]* | 259 [207; 308] | -197 [-106; -273] | 460 [174; 576]* |
| Ireland | 0 [0; 1]* | 2 [1; 4]* | -2 [0; -4] | 5 [4; 6]* | 2 [1; 2]* | 0 [0; 0] |
| Italy | 12 [7; 17]* | 5 [1; 10] | 97 [-18; 214]* | 68 [54; 81]* | 2 [1; 2] | 5 [2; 6]* |
| Lithuania | 0 [0; 0] | 1 [0; 2] | -5 [1; -10] | 0 [0; 0] | 0 [0; 1] | 0 [0; 0] |
| Macedonia | 0 [0; 0] | 0 [0; 0] | 7 [-1; 16] | 0 [0; 0] | 0 [0; 0] | 1 [0; 1] |
| Norway | 1 [0; 1]* | 0 [0; 0] | 0 [0; -1] | 6 [4; 7]* | 0 [0; 0] | 0 [0; 0] |
| Poland | 1 [1; 2] | 14 [4; 24] | -33 [6; -73] | 7 [6; 9] | 5 [3; 7] | -2 [-1; -3] |
| Portugal | 2 [1; 3]* | -1 [0; -2] | 0 [-8; 8]* | 13 [11; 16]* | 0 [0; -1] | 0 [0; 0]* |
| Slovakia | 0 [0; 0]* | 2 [1; 4]* | 11 [-2; 25]* | 1 [1; 2]* | 1 [0; 1]* | 1 [0; 1]* |
| Spain | 10 [5; 14]* | 1 [-1; 3] | -48 [-29; -68]* | 83 [65; 100]* | 1 [1; 2] | -2 [-1; -2]* |
| Switzerland | 1 [0; 1]* | 4 [1; 7]* | -10 [12; -33]* | 6 [5; 7]* | 2 [1; 3]* | -1 [0; -1]* |
| Thailand | 0 [0; 0]* | 0 [0; 0] | -14 [3; -32] | 0 [0; 0]* | 0 [0; 0] | -1 [0; -2] |
| United Kingdom | 9 [5; 13]* | 18 [12; 24] | 0 [0; 0] | 73 [58; 88]* | 5 [3; 7] | 0 [0; 0] |
| United States | 0 [0; 0]* | 0 [0; 0]* | 0 [0; 0] | 0 [0; 0]* | 0 [0; 0]* | 0 [0; 0] |

**Table S3.**
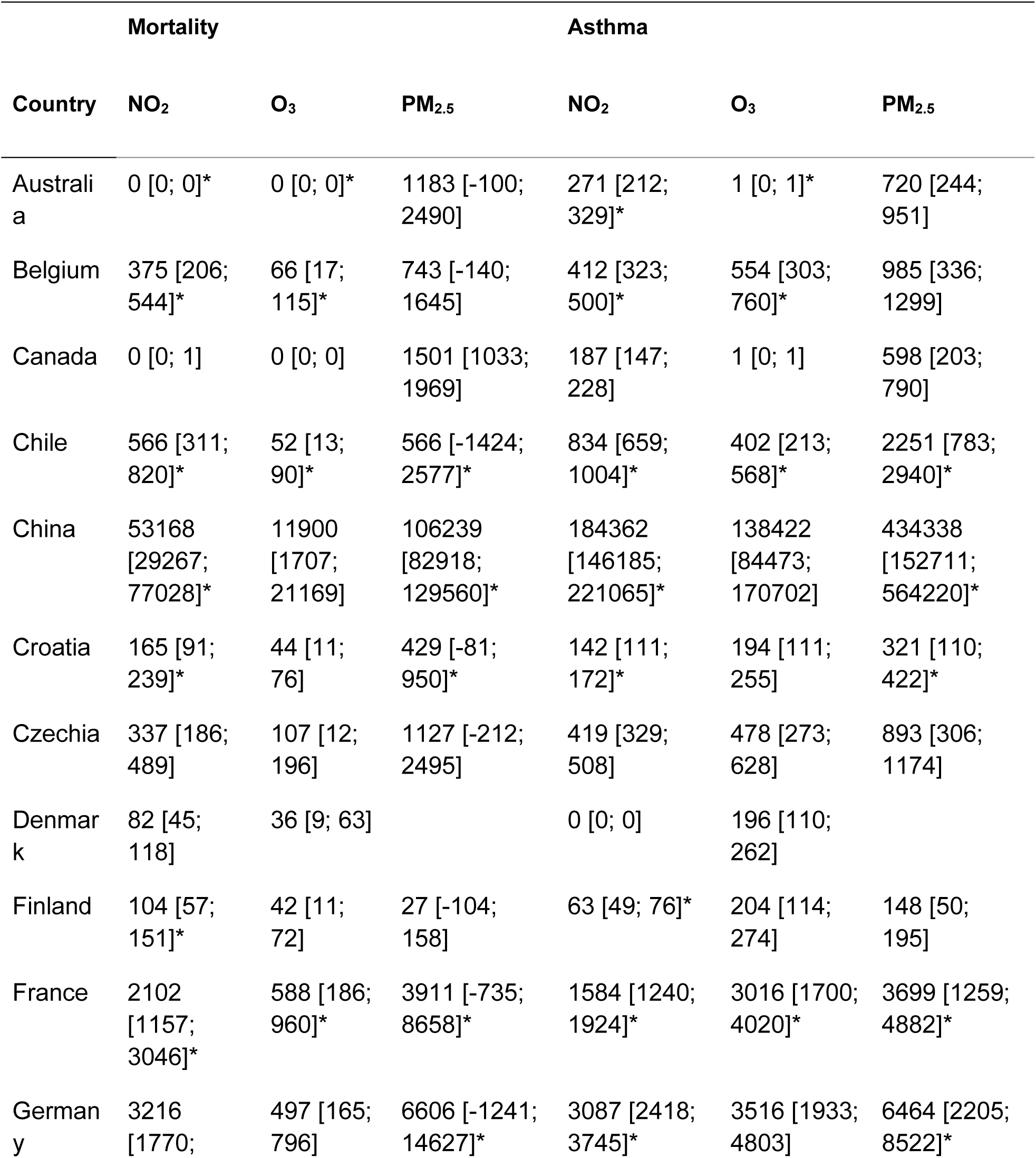
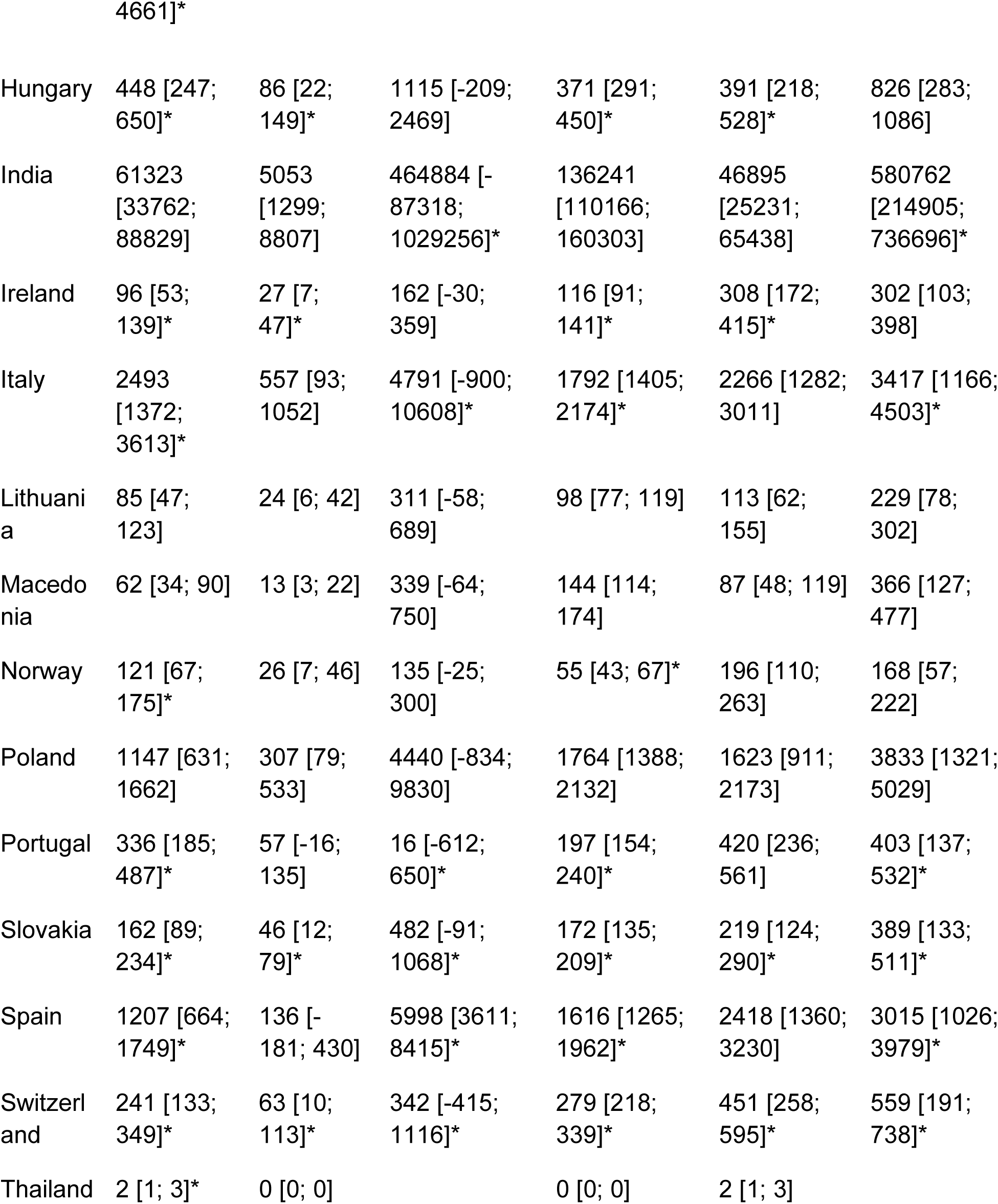
Projected health burden response. Potential premature deaths and asthma incidence that might be avoided between April and December 2020 assuming pollutant levels remain at lockdown levels (NO_2_: −29%; O_3_: −11%; PM_2.5_: −9%). Country averages and 95% confidence intervals are reported. Numbers are rounded to the nearest whole number. Values with significant (p < 0.05) pollutant anomalies after correcting for meteorological parameters are indicated with *.

